# A New Foundation Model’s Accuracy in Glaucoma Detection using Ocular Coherence Tomography Images

**DOI:** 10.1101/2024.08.04.24311475

**Authors:** Benton Chuter, Justin Huynh, Evan Walker, Shahin Hallaj, Jalil Jalili, Jeffrey Liebmann, Massimo A Fazio, Christopher A. Girkin, Robert N. Weinreb, Mark Christopher, Linda M. Zangwill

## Abstract

**Purpose:** To fine tune and evaluate the performance of the retinal foundation model (RETFound) on a diverse longitudinal clinical research dataset in glaucoma detection from optical coherence tomography (OCT) RNFL scans. Subanalyses of the model performance were evaluated across different subgroups, various dataset sample sizes and training cycles (epochs).

**Design:** Evaluation of a diagnostic technology

**Subjects, Participants, and Controls:** 15,216 Spectralis OCT RNFL circle scans of 747 individuals of diverse race (56.9% White, 37.8% Black/African American, and 5.3% Other/Not reported, glaucoma severity (30.8% mild, 18.4% moderate-to-severe, and 50.9% no glaucoma), and age (44.8% <60 years, 55.2% >60 years) from the Diagnostic Innovations in Glaucoma Study (DIGS) and the African Descent and Glaucoma Evaluation Study (ADAGES). All OCT scans were labeled as “Non-glaucomatous” or “Glaucomatous.“

**Methods:** RETFound was employed to perform binary glaucoma classification. The diagnostic accuracy of RETFound was iteratively tested across different combinations of dataset sample sizes (50 to 2000 OCT RNFL circle scans), epochs (5 to 50), and study subpopulations stratified by severity of glaucoma, age, and race).

**Main Outcome Measures:** Area under receiver operating characteristic curve (AUC) for classifying RNFL scans as “Non-glaucomatous” or “Glaucomatous.“

**Results:** Performance metrics improved with larger training datasets and more training cycles, rising from an AUC of 0.61 (50 training images and 5 epochs) to AUC 0.91 (2,000 training images and 50 epochs). Gains in performance were marginal as training size increased beyond 500 scans. Performance was similar across race for all training size and cycle number combinations: African American (AUC=0.90) vs other (AUC=0.93). RNFL scans from older patients (>60 years) led to worse performance (AUC=0.85) compared to younger patients (<60 years, AUC=0.95). Performance was significantly higher for RNFL scans from patients with moderate-to-severe glaucoma vs mild glaucoma (AUC=0.99 vs 0.88, respectively).

**Conclusions:** Good RETFound performance was observed with a relatively small sample size of images used for fine tuning and across differences in race and age. RETFound’s ability to adapt across a range of OCT training conditions and populations suggests it is a promising tool to automate glaucoma detection in a variety of use cases.

**Precis:** The study found high accuracy for glaucoma detection from OCT optic nerve head RNFL scans in a diverse study population by adapting an existing foundation model (RETFound). Performance improved with larger datasets and more training cycles, achieving an AUC of 0.91 with RNFL scans alone. Results suggest RETFound is promising for automated OCT RNFL-based glaucoma detection across demographics and training conditions.

## Introduction

Glaucoma, a leading cause of blindness worldwide, is characterized by progressive optic neuropathy that can lead to irreversible vision loss if not detected and managed early.^1,2^ Optical coherence tomography (OCT) is fundamental in the diagnosis and monitoring of glaucoma, providing high-resolution cross-sectional images of the retina essential for detecting structural abnormalities associated with various eye conditions.^3,4^ However, the reliance on expert clinicians to interpret OCT images poses both substantial clinician time burden as well as issues with inter-physician variability in assessment, demonstrating the need for reliable automated systems.^5^

The advent of artificial intelligence (AI) promises to revolutionize ophthalmology by addressing these issues to aid in the diagnosis and monitoring of glaucoma.^6,7^ Recent studies have leveraged artificial intelligence and deep learning (DL) techniques to enhance glaucoma detection using OCT images, demonstrating promising results.^8–12^ For instance, Akter et al. employed VGG16, SqueezeNet, and ResNet18 models on a dataset of 780 segmented and 780 raw TSNIT OCT B-scans, resulting in an AUC of 0.93 on test data.^13^ Another multi-institutional study developed a DL model to diagnose early-onset glaucoma using spectral-domain OCT images. Pre-trained on 4,316 OCT images from 1,371 eyes with open-angle glaucoma and 193 normal eyes, and then trained on a dataset from 94 patients with early glaucoma and 84 normal subjects, the model achieved an AUC of 0.937, outperforming random forests and support vector machine models.^14^ Moreover, another study evaluated various training strategies for DL models and explored the influence of demographic and clinical factors from the study groups on the efficacy of glaucoma detection from optic disc images. It contrasts the effectiveness of deep learning algorithms by two independent investigators in different glaucoma populations, showing optimal performance with AUCs of 0.92 for any glaucoma, 0.91 for mild glaucoma, and 0.98 for moderate-to-severe glaucoma across significant datasets like DIGS/ADAGES and the Matsue Red Cross Hospital (MRCH) datasets, involving varied patient demographics from the U.S. and Japan.^12^

However, these DL models often require large, high-quality labeled datasets for effective training, which are not universally available and are resource-intensive to produce.^12,15^ This reliance on specialist-annotated data limits their scalability and applicability in diverse clinical settings.^12,15^

Self-supervised learning (SSL) helps address this labeling issue. SSL enhances data utilization by extracting features without the need for ground truth labels, creating versatile feature representations for various applications.^16–18,19^ Using large pools of unlabeled data, this approach can train robust, generalizable models that can be adapted for a variety of tasks and can outperform supervised learning methods in classification tasks.^20,21^ This characteristic makes SSL-based models a promising approach for medical applications with limited labeled data.^22,23^

Recently, an SSL-based foundation model that was trained on a large number (>1.6 million) of ophthalmic images, RETFound, was described.^24^ A primary intended use of foundation models such as RETFound is to serve as a starting model that can then be adapted and/or fine-tuned to perform a specific task of interest without needing a restrictively large or expensive training dataset.^24^ Preliminary evaluations of RETFound show its utility across multiple diseases, tasks, and imaging modalities. However, its development and testing have been primarily confined to publicly available datasets with inconsistent image and label quality and an initial dataset from the UK. To ensure its robustness and applicability in real-world settings, it is crucial to further validate RETFound using larger, more diverse datasets from multiple regions and demographic groups.

To address this need, our research focuses on a validation study of RETFound using a comprehensive dataset of OCT images from eyes with and without glaucoma. This study assesses RETFound’s performance in detecting glaucoma using OCT optic nerve head (ONH) circle scans from our unique, diverse dataset. We explore the number of images and training iterations required for fine-tuning RETFound to achieve high accuracy in this new context. Our investigation tests RETFound’s ability to detect glaucoma using OCT images, examining the impact of varying training durations and data volumes. Additionally, we assess its generalizability across different ethnicities, ages, and stages of disease to understand its performance, variability, and applicability in glaucoma detection.

## Methods

### Data Collection

This research used OCTs from the Diagnostic Innovations in Glaucoma Study (DIGS, clinicaltrials.gov ID: NCT00221897)^25^ and the African Descent and Glaucoma Evaluation Study (ADAGES, clinicaltrials.gov ID: NCT00221923).^26^ The recruitment process and methodology were approved by the institutional review boards at each participating site, adhering to the ethical standards outlined in the Declaration of Helsinki and the Health Insurance Portability and Accountability Act. All subjects provided informed consent during recruitment. While comprehensive descriptions of these studies have been presented in earlier publications,^25,26^ the critical aspects pertinent to this work are highlighted below.

The DIGS and ADAGES studies are a joint initiative between multiple institutions: the University of California San Diego Hamilton Glaucoma Center and Viterbi Family Department of Ophthalmology, the University of Alabama at Birmingham Department of Ophthalmology, and the Columbia University Medical Center Edward S. Harkness Eye Institute. The participants in these studies are a diverse mix of individuals with African, European, and Asian heritage. The studies’ protocols involve biannual collection of OCT photographs, stereo fundus images, and visual field (VF) tests as part of their ongoing research.

This analysis included a total of 15,216 Spectralis (Heidelberg Engineering GmbH, Heidelberg, Germany) OCT images of 747 participants (1231 eyes), taken from 2008 to 2019. It included macula-centered posterior pole scans from the Glaucoma Module Premier Edition, which consisted of 61 B-scans, each with 768 A-scans, covering a 30° × 25° region. Quality assessment for the SD-OCT images was conducted by the University of California, San Diego Imaging Data Evaluation and Analysis Reading Center following standardized protocols. Any SD-OCT images with low signal quality or those containing artifacts were discarded.

VF assessments were carried out with the Humphrey Field Analyzer II, using the 24-2 test pattern and the Swedish Interactive Thresholding Algorithm standard testing algorithm. Tests with fixation losses, false-negative, or false-positive errors exceeding 33% were discarded. To gauge the severity of glaucoma damage at the time of imaging, the mean deviation (MD) from the VF test taken closest to the time of image capture, and within a year, was used for all ONH images.

### Glaucoma Labels

Ground truth glaucoma status required patients to have both repeatable glaucomatous visual field damage (GVFD) and glaucomatous optic neuropathy (GON). Eyes from patients who had neither GVFD or GON were labeled as “non-glaucomatous.” In determining GON, stereophotographs underwent review by two independent, blinded graders using a stereoscopic viewer. If the two graders disagreed, a third experienced grader adjudicated. GVFD was defined as a VF PSD (P< 0.05) and/or Glaucoma Hemifield Test outside normal limits on at least two consecutive tests. Glaucomatous eyes were further categorized into two groups based on the severity of glaucoma as indicated by 24-2 VF mean deviation (MD). Patients with an MD of −6.0 or worse decibels (dB) were classified as having “moderate-to-severe” glaucoma, while those with a VF MD better than −6.0 dB were categorized as having “mild” glaucoma.

### Image Preprocessing

SD-OCT images were resized to a uniform 224 × 224 pixel dimension to meet the input requirements for RETFound. This resolution also has proven sufficient for diagnosing primary open-angle glaucoma (POAG) in earlier experiments.^27^

### Evaluation of RETFound using OCTs for glaucoma label assignment

We assessed the practical use and performance of RETFound in detecting glaucoma from these preprocessed OCT images. We conducted comprehensive iterative testing to measure RETFound’s performance, captured as the area under the receiver operating characteristic (AUC), to predict glaucoma status. This involved training RETFound with different datasets of OCT images varying in size (50, 100, 200, 500, 1,000, 2,000) and for different numbers of training epochs (5, 10, 20, 50). Through this approach, we aimed to determine whether RETFound could achieve or exceed the performance of other deep learning models in classification tasks with relatively minimal training (fine-tuning) on smaller, labeled datasets. Additionally, we evaluated whether RETFound’s results were generalizable across differences in glaucoma severity, age, and race.

### Number of Images Variation & Dataset Split

A total of 15216 images from 747 patients were randomized into training (10708 images from 512 patients), validation (1497 images from 99 patients), and test (3011 images from 212 patients) pools using a standard 70-10-20 patient-based split (as illustrated in Figure 1). Demographic information for the entire dataset and for each of these pools is available in Table 1. RETFound was then evaluated across various dataset sizes (50, 100, 200, 500, 1,000, 2,000) and epochs (5, 10, 20, 50), totaling 24 size-epoch combinations. These specific ranges were selected after initial testing indicated they provided a comprehensive spectrum of model performance, from subpar to optimal.

**Figure 1:**
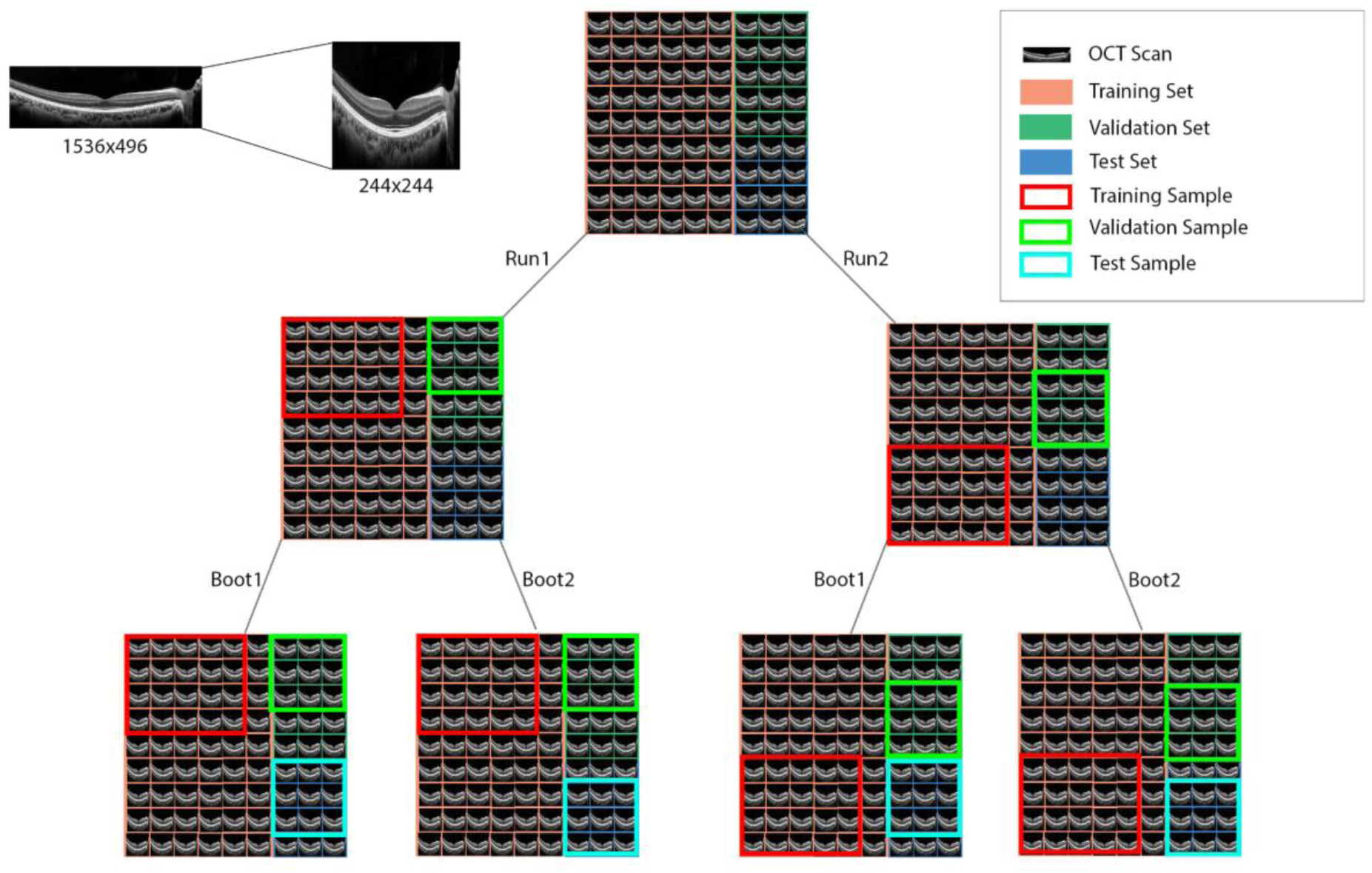
Diagram showing model workflow, involving OCT preprocessing, fine-tuning runs, & bootstraps.

**Table 1:** Overview of all Cohorts.

|  | Overall (n = 747<br>subjects; 1231 eyes;<br>15216 images) | Training (n = 512<br>subjects; 812 eyes;<br>10708 images) | Validation (n = 99<br>subjects; 167 eyes;<br>1497 images) | Testing (n = 212<br>subjects; 356 eyes;<br>3011 images) |
| --- | --- | --- | --- | --- |
| Age | 60.3 (59.2, 61.4) | 61.7 (60.4, 63.0) | 54.8 (51.4, 58.2) | 58.1 (56.1, 60.1) |
| Age Classification |  |  |  |  |
| Age < 60 | 335 (44.8%) | 205 (40.0%) | 60 (60.6%) | 114 (53.8%) |
| Age > 60 | 412 (55.2%) | 307 (60.0%) | 39 (39.4%) | 98 (46.2%) |
| Sex |  |  |  |  |
| Female | 438 (58.6%) | 282 (55.1%) | 65 (65.7%) | 134 (63.2%) |
| Male | 309 (41.4%) | 230 (44.9%) | 34 (34.3%) | 78 (36.8%) |
| Race |  |  |  |  |
| American Indian/ Alaska Native | 2 (0.3%) | 1 (0.2%) | 0 (0.0%) | 1 (0.5%) |
| Asian | 29 (3.9%) | 23 (4.5%) | 3 (3.0%) | 3 (1.4%) |
| Black or African American | 282 (37.8%) | 185 (36.1%) | 31 (31.3%) | 104 (49.1%) |
| Native Hawaiian or Other Pacific Islander | 3 (0.4%) | 3 (0.6%) | 0 (0.0%) | 0 (0.0%) |
| Unknown or Not Reported | 6 (0.8%) | 4 (0.8%) | 2 (2.0%) | 0 (0.0%) |
| White | 425 (56.9%) | 296 (57.8%) | 63 (63.6%) | 104 (49.1%) |
| Ethnicity |  |  |  |  |
| Hispanic | 18 (2.4%) | 13 (2.5%) | 5 (5.1%) | 3 (1.4%) |
| Not Hispanic | 647 (86.6%) | 446 (87.1%) | 82 (82.8%) | 181 (85.4%) |
| Unknown or Not Reported | 82 (11.0%) | 53 (10.4%) | 12 (12.1%) | 28 (13.2%) |
| Diabetes |  |  |  |  |
| No | 652 (87.3%) | 437 (85.4%) | 93 (93.9%) | 189 (89.2%) |
| Yes | 95 (12.7%) | 75 (14.6%) | 6 (6.1%) | 23 (10.8%) |
| Hypertension |  |  |  |  |
| No | 463 (62.0%) | 299 (58.4%) | 68 (68.7%) | 142 (67.0%) |
| Yes | 284 (38.0%) | 213 (41.6%) | 31 (31.3%) | 70 (33.0%) |
| 24-2 VF MD (dB) | -3.31 (-3.71, -2.92) | -4.10 (-4.62, -3.59) | -1.61 (-2.42, -0.80) | -1.08 (-1.54, -0.63) |
| Baseline Disease Severity |  |  |  |  |
| Mild Glaucoma | 379 (30.8%) | 313 (37.9%) | 25 (15.0%) | 41 (11.5%) |
| Moderate-to-severe Glaucoma | 226 (18.4%) | 195 (23.6%) | 13 (7.8%) | 18 (5.1%) |
| Non-Glaucomatous | 626 (50.9%) | 317 (38.4%) | 129 (77.2%) | 297 (83.4%) |
| Axial Length (mm) | 23.9 (23.9, 24.0) | 24.0 (23.9, 24.1) | 23.7 (23.5, 24.0) | 23.8 (23.7, 24.0) |
| Spherical Equivalent | -0.51 (-0.64, -0.38) | -0.58 (-0.74, -0.42) | -0.58 (-0.90, -0.26) | -0.31 (-0.55, -0.07) |
| IOP (mmHg) | 14.7 (14.4, 14.9) | 14.7 (14.4, 15.1) | 14.6 (14.1, 15.2) | 14.6 (14.2, 15.0) |
| CCT (μm) | 539 (536, 542) | 539 (535, 542) | 544 (536, 552) | 538 (533, 543) |
| Baseline Visit Glaucoma Classification |  |  |  |  |
| GVFD & GON | 605 (49.1%) | 508 (61.6%) | 38 (22.8%) | 59 (16.6%) |
| Non-glaucomatous | 626 (50.9%) | 317 (38.4%) | 129 (77.2%) | 297 (83.4%) |
| Last Visit Glaucoma Classification |  |  |  |  |
| GVFD & GON | 605 (49.1%) | 508 (61.6%) | 38 (22.8%) | 59 (16.6%) |
| Non-glaucomatous | 626 (50.9%) | 317 (38.4%) | 129 (77.2%) | 297 (83.4%) |

IOP: intraocular pressure, CCT: central corneal thickness, VF: visual field, MD: mean deviation
\*Number of patients when stratified by characteristic may not sum to total (2104 for “All”); this remainder were unreported for the characteristic of interest.
\*\*Age of 5 subjects progressed past 60y during the study; these are here reported at baseline age <60y.

For each of these size-epoch combinations, models were trained, validated, and tested on subsets randomly sampled from the predetermined training, validation, and test pools in a 70-10-20 ratio. The total number of images reported represented the current size for the specific size-epoch combination being evaluated (number of training samples + number of validation samples = current size), as shown in Figure 1. To account for and assess variability across different training runs, each size-epoch combination underwent 10 separate training runs, with the sampling process repeated for each. For each of these training runs, an additional 100 bootstrap runs were performed, resulting in a total of 240 training runs and 24,000 bootstrap runs. Further implementation details are provided in Supplementary Table 2.

### Analysis

Given the balanced distribution of glaucoma cases within the DIGS/ADAGES dataset (as outlined in Table 1), each run’s performance was measured using the area under the receiver operating characteristic (AUC). 95% confidence intervals (CIs) for these were calculated using a cumulative density function, sorting the bootstrapped estimates and selecting the values corresponding to the 2.5th and 97.5th percentiles to determine the CI range. This approach effectively captures variability between runs, especially when the image count is high. The generalizability of the models was also assessed by stratifying the results by race (Black vs. non-Black), age (<60 years vs. ≥60 years), and severity of glaucoma (MD >-6.0 dB vs. MD ≤-6.0 dB)).

## Results

This study included 15216 images from 747 subjects and 1232 eyes, divided into subsets for testing (3011 images from 212 subjects and 356 eyes), training (10708 images from 512 subjects and 812 eyes), and validation (1497 images from 99 subjects and 167 eyes), as shown in Table 1. The average age of participants in the study is 60.3 years, with 44.8% (n=335) under 60 years of age and 55.2% (n=412) over 60 years. Females (n=438, 58.6%) outnumber males (n=309, 41.4%). The majority of the study population is White (56.9%, n=425), followed by Black (37.8%, n=282) and Asian (3.9%, n=29) individuals. The racial status for 6 (0.8%) participants was unspecified or unrecorded. Eye-level characteristics such as VF MD, axial length, spherical equivalent, intraocular pressure (IOP), and central corneal thickness (CCT) are documented for the training, validation, and test sets in Table 1. 30.8% (n=379) of patients’ eyes indicated mild glaucoma (VF MD >-6 dB) compared to 18.4% (n=226) with moderate-to-severe glaucoma (VF MD <-6 dB), while 50.9% (n=626) had no glaucoma. The datasets are evenly distributed between the two outcomes of interest at the latest visit: Glaucoma (n=605, 49.1%) and Not Glaucoma (n=626, 50.9%).

Table 2, Figure 2, and Supplementary Figures S1 and S2 demonstrate the model’s performance across various epoch and OCT image sample size combinations. Increases in either epoch count or sample size were associated with increased performance, however, sample size had a larger impact than epoch count. Across all epochs, as the image count grew from 50 to 2000, the AUC also increased; at a constant of 50 epochs, this increase ranges from 0.64 at 50 images to 0.85 at 200 images and 0.91 at 2000 images. At a constant sample size of 2,000 images, AUC increased from 0.86 at 5 epochs to 0.91 at 50 epochs. The rate of improvement diminished as the sample size increased, with large gains in performance when moving from 50 to 500 images and relatively small gains when adding additional images after 500.

**Table 2:**
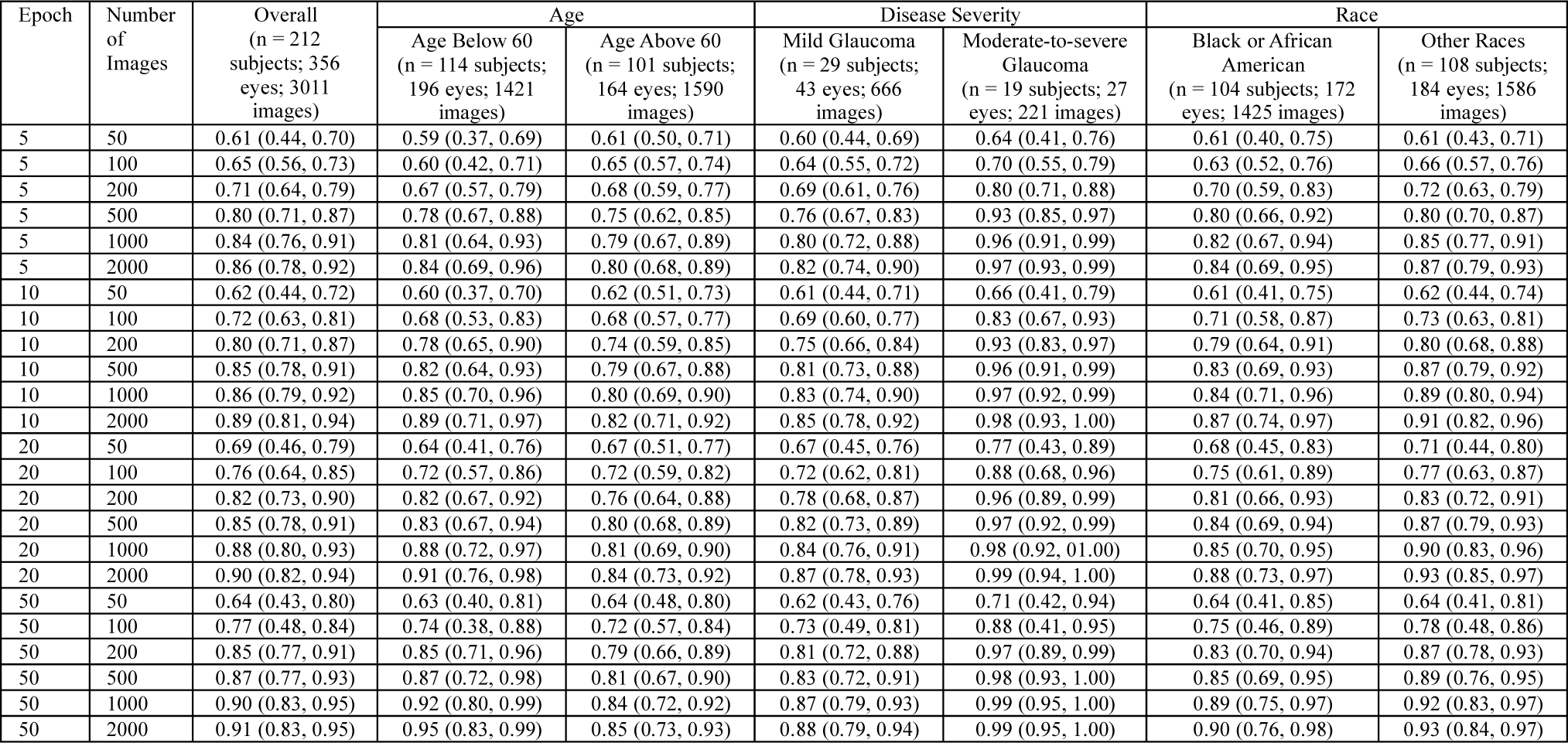
Summary of the model performance on the local datasets as captured by AUC, with 95% confidence intervals. The number of images represents the sum of images used for training and validation. Stratified by Age, Disease Severity, and Race.

**Figure 2:**
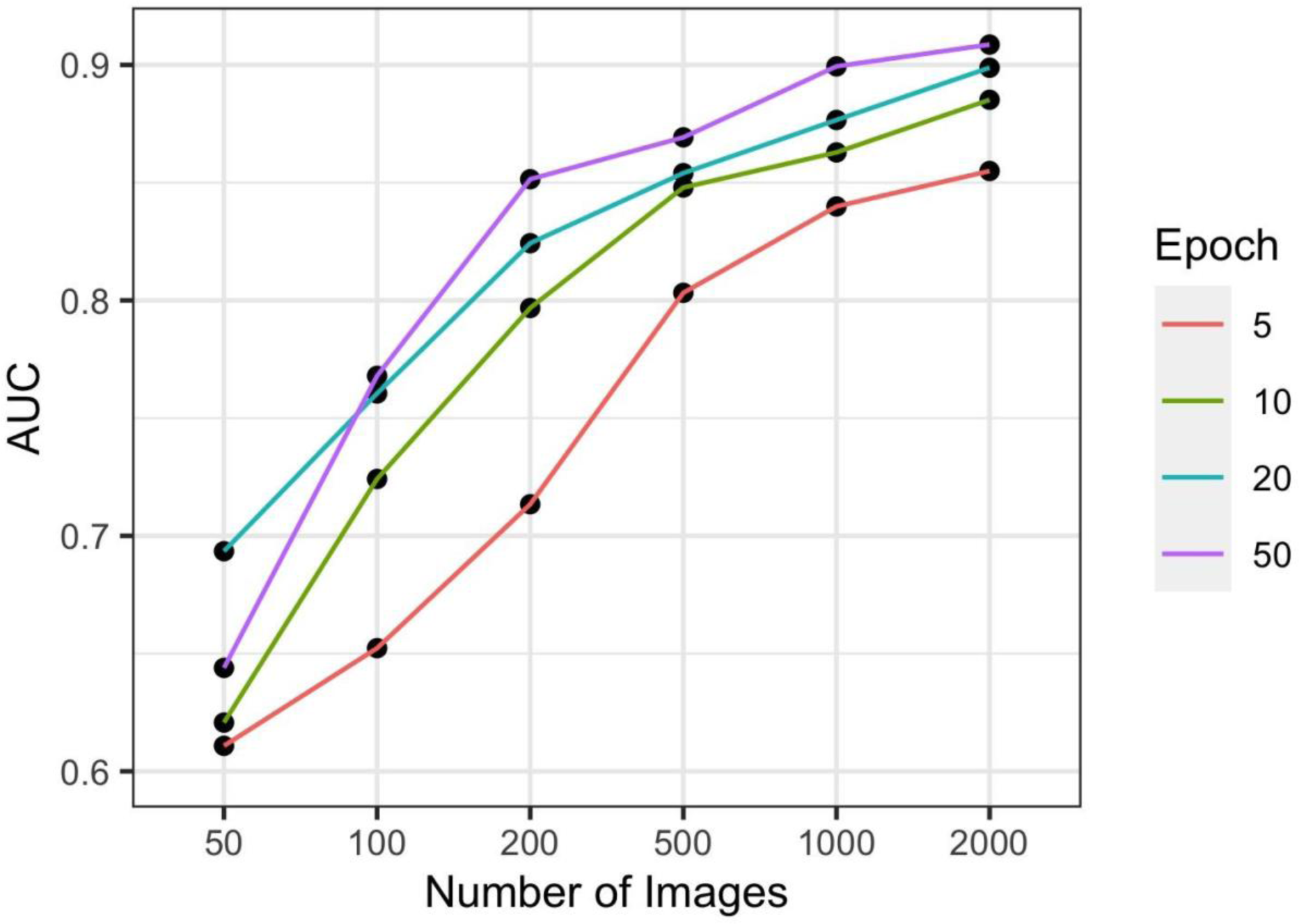
Plots demonstrating the relationship between number of images (x) vs and performance, as measured by Area under the receiver operating characteristic curve (AUC) (y), at each tested epoch number.

The 95% confidence intervals for AUC narrow with the increase in the number of images, indicative of higher confidence in the AUC values with larger datasets; at a constant 50 epochs, the 95% CI range is 0.37 at 50 epochs, 50 images and decreases to 0.12 at 50 epochs, 2000 images. This narrowing of 95% CIs with the increased sample size and epoch count indicates better model stability and performance consistency, as more data is available for training over greater numbers of epochs. An increase in the number of images appears to have a larger effect than a proportional increase in epochs. For 2000 training and validation images, the CI range only decreases from 0.14 at 5 epochs to 0.12 at 50 epochs. Model performance at 2000 images, over 50 epochs, consistently achieves excellent performance with a mean AUC of 0.91.

Model performance was also assessed in relation to demographic factors (Table 2, Figure 3 and Supplemental Figure 3) across sample sizes and epoch numbers. With respect to age groups (<60 years vs. ≥60 years), AUCs were comparable at small sample sizes, but AUC was consistently higher in the <60 years group at larger sample sizes (0.95 (0.83 – 0.99) vs. 0.85 (0.73 – 0.93) at 2,000 images and 50 epochs). These differences were not statistically significant. Similarly in comparing racial groups (Black / African American vs. White), results were comparable at small sample sizes, but AUC slightly in the White participants at larger sample sizes (0.93 (0.84 – 0.97) vs. 0.90 (0.76 – 0.98) at 2,000 images and 50 epochs). Again, these differences were not statistically significant. Finally, with respect to disease severity, AUC was consistently higher for the moderate-to-severe group than the early glaucoma group (0.95 (0.83 – 0.99) vs. 0.85 (0.73 – 0.93) at 2,000 images and 50 epochs). These results were statistically significant at the 500, 1,000, and 2,000 sample size cases across all epoch numbers (0.93 (0.84 – 0.97) vs. 0.90 (0.76 – 0.98) at 2,000 images and 50 epochs).

**Figure 3:**
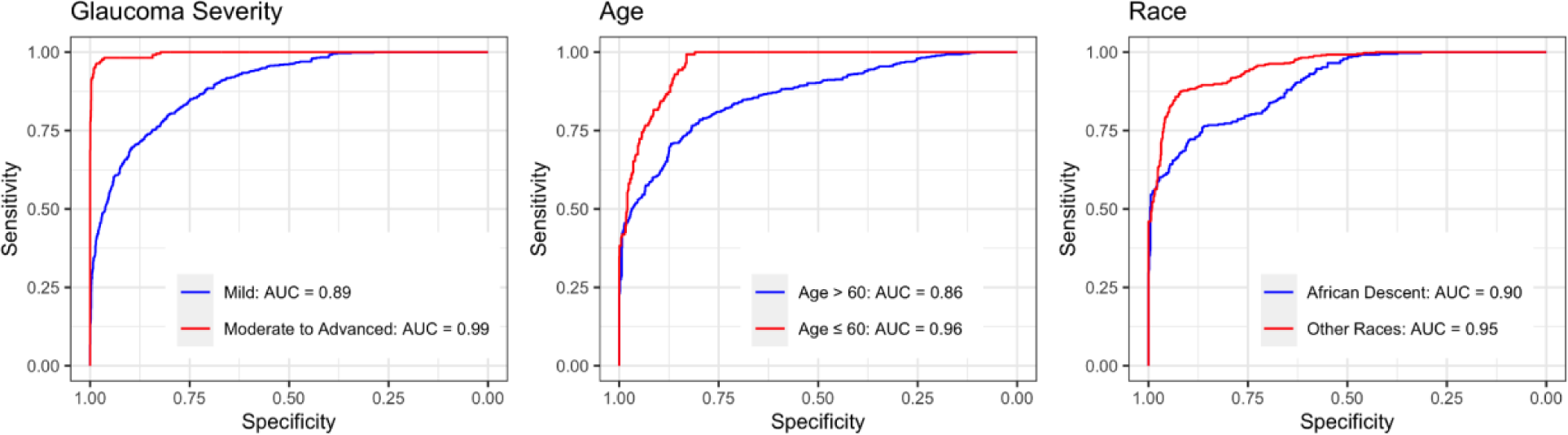
Best performing RETFound models for age, race, and severity of glaucoma. Area under the receiver operating characteristic curve (AUC) curves are stratified by Glaucoma Severity (left: Mild, Moderate-to-severe), Age (middle: >60y, <60y) and Race (right: African Descent, Other). The best performing model was defined as the model with the highest combined (AUC), fine-tuned from a single training run with 2000 images.

## Discussion

RETFound demonstrates strong diagnostic performance for OCT images, benefiting from larger datasets and longer training times. Its efficiency and accuracy make it a promising tool for clinical applications. The performance of RETFound varied with the number of images and epochs during training and was generalizable across differences in age and race. As the number of images and epochs increases, there is a general trend of improvement in diagnostic accuracy, with limited improvement in diagnostic accuracy after increasing the sample size from 500 (average AUC 0.87, 25.0 patients, 40.1 eyes) to 1000 (average AUC 0.90, 50.0 patients, 80.2 eyes) images. This suggests that although RETFound benefits from a larger volume of data and extended training, good diagnostic performance is possible with relatively small sample sizes. Likely because this foundation model is pre-trained on a large dataset using a self-supervised approach, it is able to acquire strong prior knowledge of informative retinal image features, allowing for efficient fine-tuning using smaller sample sizes and fewer epochs to achieve strong performance.

RETFound matches or exceeds the performance of previously developed convolutional neural networks (CNNs) based DL methods while requiring significantly fewer samples for training.^28^ When trained for 50 epochs on 200 images, it begins to approach the performance of ResNet-50, which was trained on much larger DIGS/ADAGES OCT datasets comprising tens of thousands of images while using a similar definition of glaucoma (GVFD and GON) as well as comparable distribution of severity of disease.^28^ Specifically, RETFound achieves an AUC of 0.87 (95% CI: 0.77–0.93) with just 500 images, matching the performance of the prior CNN (AUC = 0.86, 95% CI: 0.84–0.87, n=25,751).^28^ Additionally, RETFound surpasses previous CNNs when trained and validated on more than 500 images, achieving an AUC of 0.90 (95% CI: 0.83–0.95) for 1000 images. Even with only 20 epochs of training, RETFound exceeds the performance of prior CNNs with an AUC of 0.88 (95% CI: 0.80–0.93) when trained on 1000 OCT images. RETFound OCT also shows comparable if not superior performance to previous CNNs using ONH fundus images. Specifically, RETFound OCT achieved an AUC of 0.91 (95% CI: 0.83–0.95) when trained and validated on only 2000 OCT images, similar to the AUC of 0.91 (95% CI: 0.89–0.92) reported for CNNs trained on 20,828 ONH fundus images.^28^

RETFound’s performance also compares favorably to previous transformer models applied to the DIGS/ADAGES dataset. While a transformer model achieved an AUC of 0.92 on 22,464 OCT images from the DIGS/ADAGES dataset,^29^ RETFound achieved similar results with an AUC of 0.91 (95% CI: 0.83–0.95) using just one-tenth of the data. This outcome demonstrates RETFound’s efficiency, as it requires fewer labeled training samples to match or surpass the performance of prior approaches, benefiting from both increased training time and sample size.

In the original study,^24^ RETFound’s application to a publicly available dataset for glaucoma classification yielded comparable or mildly inferior outcomes compared to its performance following fine-tuning on the clinical DIGS/ADAGES dataset. This includes glaucoma detection on the PAPILA dataset, for which they reported a mean AUC of 0.86 (0.84, 0.87).^24^ However, it should be noted that the original study did not directly test RETFound’s performance with OCT for glaucoma prediction, only fundus images for Glaucoma and “Multi-class disease”, such that a direct comparison cannot be cleanly made.

Further, discrepancies in performance may result from various factors, including variations in ground truth definition of glaucoma disease severity, study population, image quality, or other elements.^30^ Numerous studies have reported high accuracy in glaucoma detection; however, direct comparisons across studies can be difficult due to data source differences, including variations in disease severity, which is often not reported despite its significant impact on accuracy.^7,31^ In particular, accuracy for identifying mild glaucoma is often substantially lower than identifying moderate-to-severe disease,^12,32^ as shown in this work, where mean AUC for detecting moderate-or-severe glaucomatous disease rose to 0.99 (0.95, 1.00) at 2000 images and 50 epochs, compared to 0.88 (0.79, 0.94) for detecting mild disease.

When comparing RETFound model performance when stratified for detecting mild versus moderate-to-severe glaucoma, significant differences are observed. At 50 epochs, the model’s performance on 1000 images from patients with mild glaucoma shows a mean AUC of 0.87 (95% CI: 0.787 to 0.934), whereas for moderate to severe glaucoma, the mean AUC significantly improves to 0.986 (95% CI: 0.950 to 0.999). Similarly, with 2000 images, the AUC for mild glaucoma is 0.881 (95% CI: 0.791 to 0.937), compared to an AUC of 0.991 (95% CI: 0.954 to 0.999) for moderate to severe cases. Notably, even with just 200 images, the model achieved an impressive AUC of 0.965 (95% CI: 0.887 to 0.992) for moderate to severe glaucoma. These findings highlight the model’s enhanced sensitivity in detecting more advanced stages of glaucoma. This may be attributed to the more pronounced structural changes in the optic nerve head and retinal nerve fiber layer in moderate-to-severe glaucoma, which are more easily recognized by the model. The subtle changes in mild glaucoma may present a greater challenge for detection, requiring higher image resolution or additional clinical features to improve model performance.

This study also finds that the RETFound model maintains strong performance across various demographic groups, showing no statistically significant differences in AUC when stratified by race. This outcome implies that the model effectively generalizes across diverse populations, a crucial trait for its clinical use in different real-world scenarios. However, while not reaching statistical significance, our results suggest that the RETFound model’s performance may vary between age groups. Specifically, when trained for 50 epochs on 2000 images the model achieved an AUC of 0.95 (95% CI: 0.83 to 0.99) for subjects below 60 years of age, whereas for those above 60, the AUC was lower at 0.85 (95% CI: 0.73 to 0.93). This may be due in part to differences in the severity of disease among these populations. This suggests that the model is more effective at detecting glaucoma in younger patients, potentially due to more pronounced retinal changes in younger individuals or differences in disease pathology. The ROC curve in Figure 3 further illustrates these differences, with the curve for subjects under 60 showing higher sensitivity and specificity across most thresholds compared to those over 60. This discrepancy could be due to age-related changes in retinal structures that are harder to detect in older individuals or reflect differences in the underlying disease pathology. These findings underscore the importance of considering demographic factors such as age in the development and evaluation of AI models for medical diagnostics. While the RETFound model shows promise, its varying performance across age groups highlights the need for further refinement and possibly the development of age-specific models or adjustments ^11^.

One limitation is that this study did not explicitly explore the impact of batch size on training dynamics, as theoretically batch size could affect gradient smoothness and convergence stability. However, initial experiments showed no significant effect on results, and relevant hyperparameters are included in Supplemental Table 2. The methodology of this study is also limited by its binary classification approach, distinguishing only between glaucoma and non-glaucoma. This simplification may not adequately capture the nuanced spectrum of ocular diseases and the variability in normal human optic nerve head structure. Enhanced diagnostic accuracy in stratifying disease severity suggests that a model with a broader range of categories that includes glaucoma suspects, or glaucomatous optic nerve damage without visual field damage, could be more beneficial for glaucoma detection.^24^ However, a binary classification system is essential for generating referral suggestions and plays a key role in telehealth, screening, primary care, and clinical decision-making tools. Relying solely on OCT RNFL imaging also has its limitations, and incorporating additional imaging techniques or diagnostic information could enhance the model’s performance. Additionally, using a relatively uniform dataset in this study may limit the applicability of the results to diverse populations. These limitations are not unique to glaucoma detection and reflect broader challenges often faced when implementing AI-driven methods in ophthalmology.

Overall, the findings underscore RETFound’s adaptability and efficiency across various training configurations, demonstrating significant performance gains even with limited training samples or computational resources—common constraints in real-world clinical settings. Many healthcare facilities face challenges in acquiring large volumes of expertly labeled data and the necessary computational infrastructure for extensive model training. Models developed externally on separate data often may suffer worse performance when applied to an independent local dataset. As such, RETFound’s reduced dependence on extensive labeled datasets and its ability to maintain high performance across diverse training conditions make it a viable and innovative tool for integrating AI into ophthalmological practices. Fine-tuning enables models to be adapted to specific clinical settings, patient demographics, and disease presentations, thereby optimizing their diagnostic accuracy and utility. Crucially, models can reach high performance with small dataset sizes; this study suggests fine-tuning with only 500 or even 200 images may be sufficient for accurate glaucoma detection. This study highlights the potential of foundational models trained on large, unlabeled datasets to overcome barriers to AI adoption, enhancing glaucoma detection in telehealth, primary care, community, and clinical environments.

Future efforts will focus on integrating fundus images as well as OCT data in models to enable a multimodal approach. By combining fundus and OCT imaging for multimodal assessment of RETFound, its diagnostic potential can be further evaluated and performance may be improved. Expanding the validation study to cover a broader spectrum of eye conditions, beyond just glaucoma, to include multiple disease categories, could also greatly enhance our understanding of RETFound’s flexibility and significance. Foundational AI models have the potential to significantly advance the field of ophthalmology, but they require extensive validation before they can be implemented in clinical practice. Furthermore, including diverse and representative datasets in these studies will be essential for evaluating the models’ real-world performance and reliability.

## Data Availability

Full results described in the present study are available upon reasonable request to the authors. Public access to images and associated clinical data sets is limited.

## Supplemental Figures

**Supplemental Figure 1:**
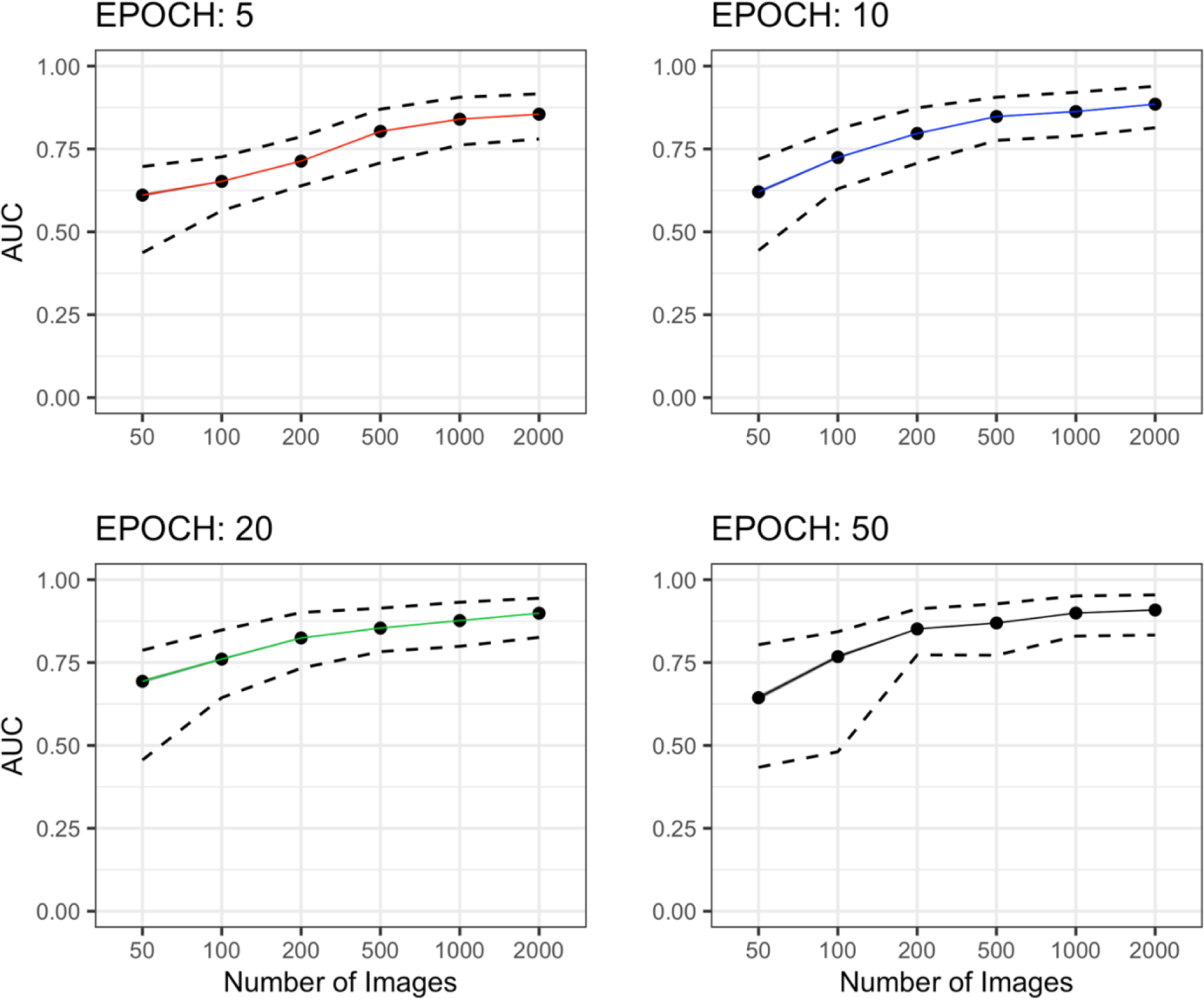
Plots demonstrating the relationship between number of images (x) vs and performance, as measured by AUC (y), at each tested epoch number. Dashed lines correspond to the range of 95% Cis.

**Supplemental Figure 2:**
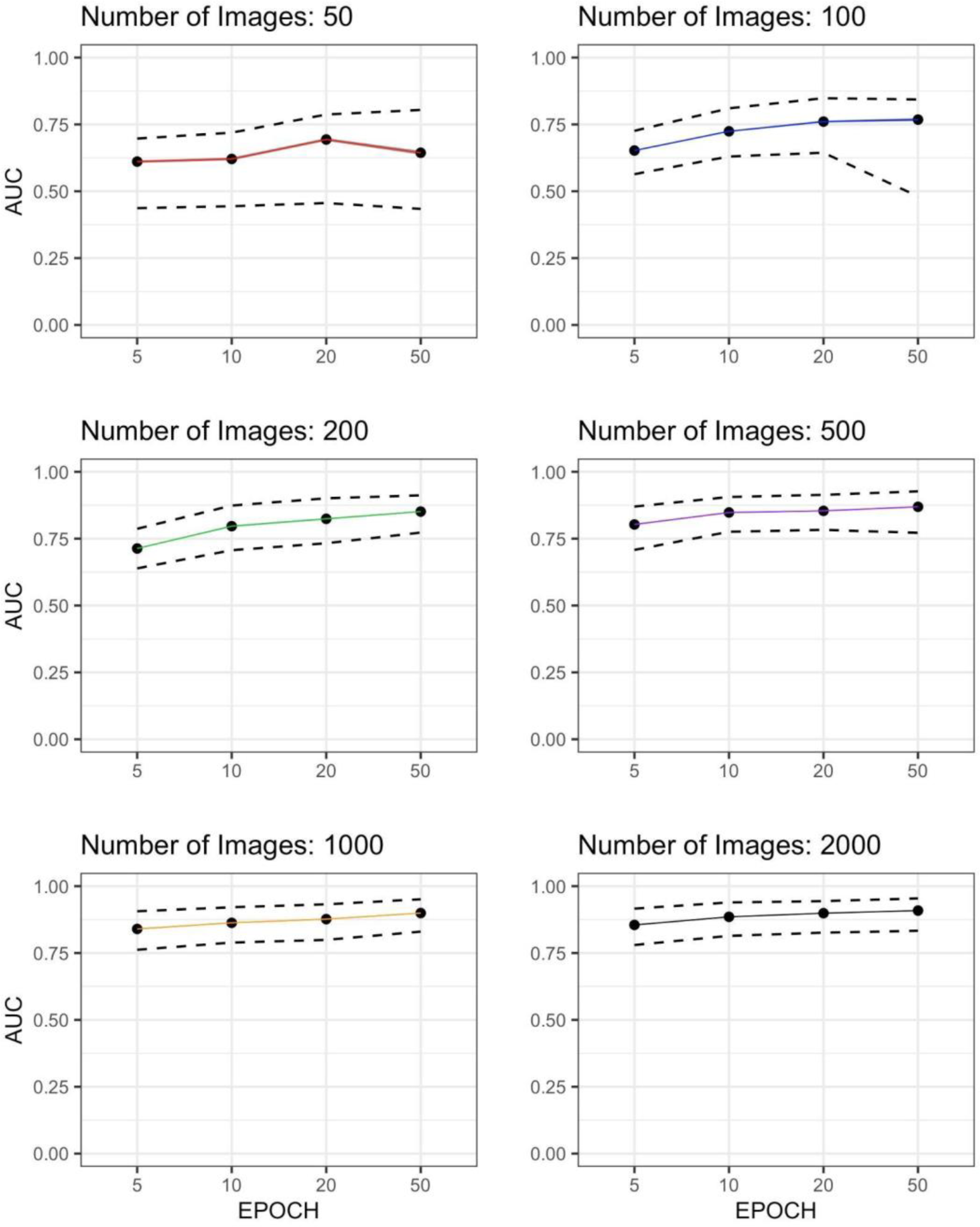
Plots of the relationship between number of epochs (x) and diagnostic performance, as measured by area under the receiver operating characteristic curve (AUC, y), at each tested dataset sample size. Dashed lines correspond to the range of 95% Cis.

**Supplemental Figure 3:**
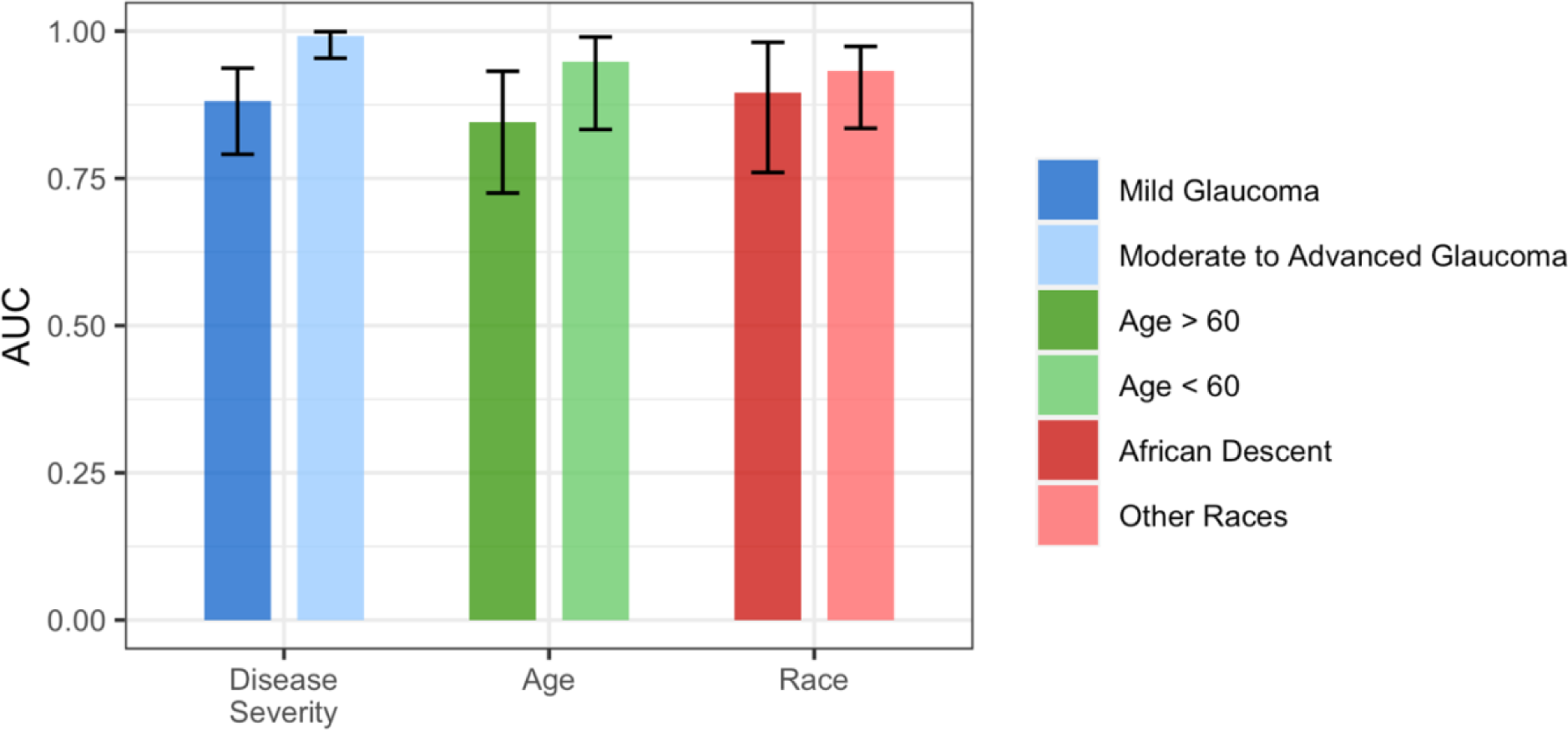
Bar Plots of mean area under the receiver operating characteristic curve (AUC) values for 50 epochs, 2000 training/validation images Stratified by Age (<60 years, ≥60 years), glaucoma severity (Mild, Moderate-to-severe) and race (African Descent, Other), with confidence intervals.

## Supplemental Tables

**Supplemental Table 1.**
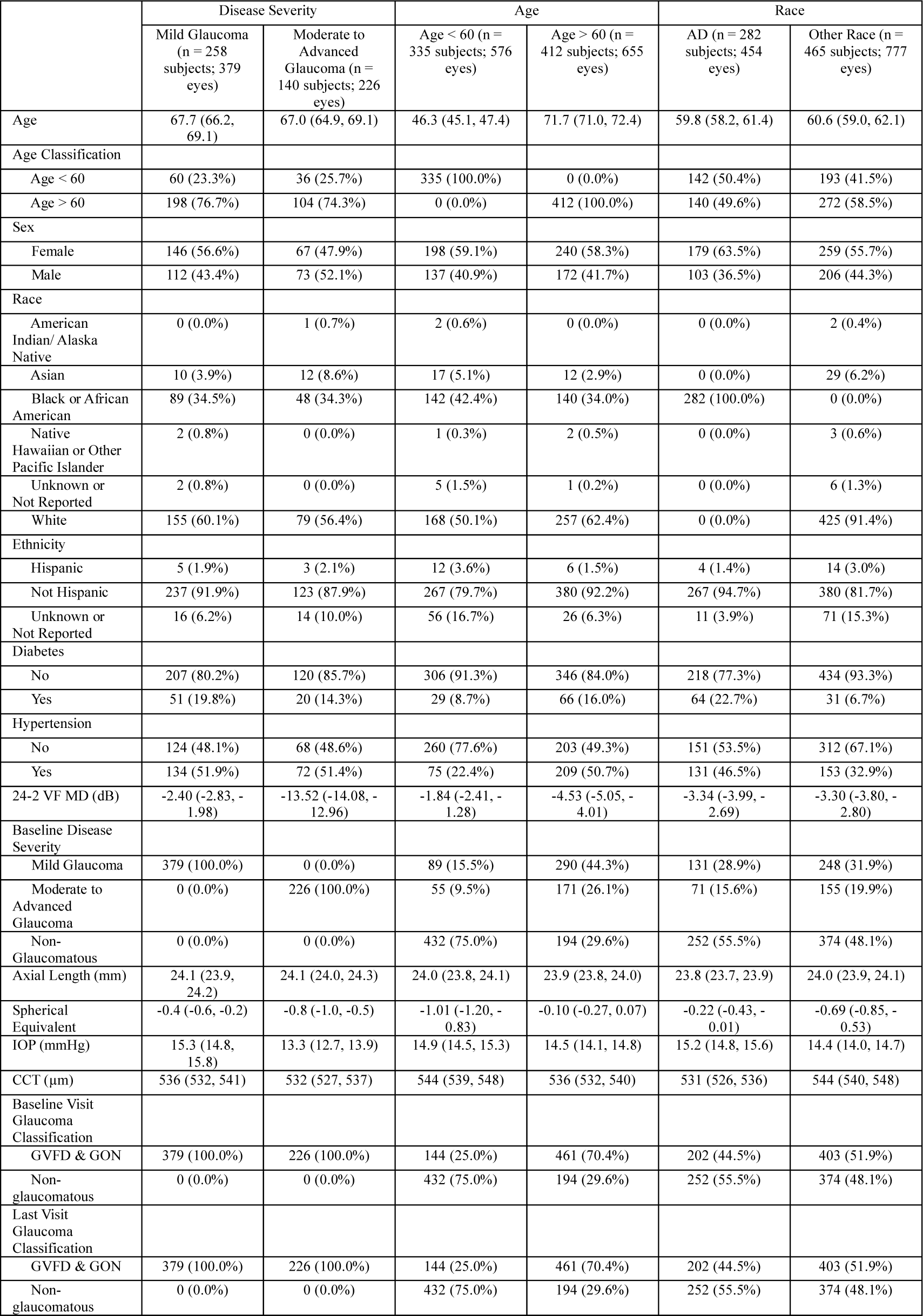

**Supplemental Table 2.**

| Parameter | Train Command Value |
| --- | --- |
| Processing Unit | A40 |
| Mode | Training |
| Optimizer | AdamW |
| --nproc_per_node | 1 |
| --batch_size | 16 |
| --world_size | 1 |
| --model | vit_large_patch16 |
| --epochs | EPOCH |
| --blr | 5e-3 |
| --layer_decay | 0.65 |
| --weight_decay | 0.05 |
| --drop_path | 0.2 |
| --nb_classes | 2 |
| --input_size | 244 |

## Notes

### Competing Interest Statement

Benton Chuter and Justin Huynh report no conflicts, with Huynh only listing the T35 grant from the National Research Service Award (NRSA). Linda M. Zangwill discloses support from the National Eye Institute, The Glaucoma Foundation, National Institutes of Health, and Heidelberg Engineering, along with consulting fees from AbbVie and Topcon Medical Systems, and involvement with AISight Health Inc. Evan Walker lists grants from the National Eye Institute and P30EY022589. Christopher A. Girkin discloses funding from the National Eye Institute, Topcon, EyeSight Foundation of Alabama, Research to Prevent Blindness, and Heidelberg Engineering. Mark Christopher and Robert N. Weinreb also acknowledge support from the National Eye Institute and other institutions, with Weinreb additionally reporting equipment support from Centervue, Optovue, and Topcon Medical. Massimo Fazio details funding from the National Eye Institute, EyeSight Foundation of Alabama, Research to Prevent Blindness, Heidelberg Engineering GmbH, and Topcon Healthcare Inc., with research support from Wolfram Research Inc. Shahin Hallaj reports a stipend from the NIH Bridge2AI common fund. Jalil Jalili discloses no conflicts.

### Funding Statement

Benton Chuter lists support from UCSD MEDGAP 2023. Linda M. Zangwill discloses funding from the National Eye Institute, The Glaucoma Foundation, National Institutes of Health, and Heidelberg Engineering. Evan Walker lists grants from the National Eye Institute and P30EY022589. Christopher A. Girkin discloses support from the National Eye Institute, Topcon, EyeSight Foundation of Alabama, Research to Prevent Blindness, and Heidelberg Engineering. Mark Christopher acknowledges funding from the National Eye Institute, The Glaucoma Foundation, K99EY030942, and R00EY030942. Massimo Fazio details support from the National Eye Institute, EyeSight Foundation of Alabama, Research to Prevent Blindness, Heidelberg Engineering GmbH, Topcon Healthcare Inc., and research support from Wolfram Research Inc. Shahin Hallaj reports a stipend from the NIH Bridge2AI common fund. Robert N. Weinreb discloses funding from Topcon Medical, the National Institutes of Health, Research to Prevent Blindness, and equipment support from Centervue, Optovue, and Topcon Medical.

### Author Declarations

The recruitment process and methodology were approved by the institutional review boards at each participating site, adhering to the ethical standards outlined in the Declaration of Helsinki and the Health Insurance Portability and Accountability Act. All subjects provided informed consent during recruitment. IRB of The University of California San Diego Hamilton Glaucoma Center and Viterbi Family Department of Ophthalmology gave ethical approval for this work. IRB of The University of Alabama at Birmingham Department of Ophthalmology gave ethical approval for this work. IRB of The Columbia University Medical Center Edward S. Harkness Eye Institute gave ethical approval for this work.

